# Evaluating prediction of short-term tolerability of five type 2 diabetes drug classes using routine clinical features: UK population-based study

**DOI:** 10.1101/2025.02.17.25322406

**Authors:** Pedro Cardoso, Katie G. Young, Rhian Hopkins, Bilal A. Mateen, Ewan R. Pearson, Andrew T. Hattersley, Trevelyan J. McKinley, Beverley M. Shields, John M. Dennis, the MASTERMIND consortium

## Abstract

**Introduction:** A precision medicine approach in type 2 diabetes (T2D) needs to consider potential treatment risks alongside established benefits for glycaemic and cardiometabolic outcomes. Considering five major T2D drug classes, we aimed to describe variation in short-term discontinuation (a proxy of overall tolerability) by drug and patient routine clinical features and determine whether combining features in a model to predict drug class-specific tolerability has clinical utility.

**Methods:** UK routine clinical data (Clinical Practice Research Datalink, 2014–2020) of people with T2D initiating glucagon-like peptide-1 receptor agonists (GLP-1RA); dipeptidyl peptidase-4 inhibitors (DPP4i); sodium-glucose co-transporter-2 inhibitors (SGLT2i); thiazolidinediones (TZD) and sulfonylureas (SU) in primary care were studied. We first described the proportions of short-term (3-month) discontinuation by drug class across subgroups stratified by routine clinical features. We then assessed the performance of combining features to predict discontinuation by drug class using a flexible machine learning algorithm (Bayesian Additive Regression Trees).

**Results:** Amongst 182,194 treatment initiations, discontinuation varied modestly by clinical features. Higher discontinuation on SGLT2i and GLP-1RA was seen for older patients and those with longer diabetes duration. For most other features, discontinuation differences were similar by drug class, with higher discontinuation for patients who had previously discontinued metformin, females, and people of South-Asian and Black ethnicities. Lower discontinuation was seen for patients currently taking statins and blood pressure medication. The model combining all sociodemographic and clinical features had a low ability to predict discontinuation (AUC = 0.61).

**Conclusion:** A model-based approach to predict drug-specific discontinuation for individual patients with T2D has low clinical utility. Instead of likely tolerability, prescribing decisions in T2D should focus on drug-specific side-effect risks and differences in the glycaemic and cardiometabolic benefits of available medication classes.

**Key message:** Routine clinical features are not sufficient to predict individuals likely to discontinue (maybe say tolerate due to the proxy stuff) T2D glucose treatment.

**Why did we undertake this study?:** Precision medicine studies aiming to guide the choice of type 2 diabetes treatment have mainly evaluated treatment benefits, meaning there is little evidence to inform targeting based on potential treatment risks.

**What is the specific question(s) we wanted to answer?:** Can routine clinical features be used to predict individual patients likely to discontinue therapy short-term (a proxy of overall drug tolerability), considering five major type 2 diabetes glucose-lowering drug classes.

**What did we find?:** Routine clinical features are associated with differences in short-term tolerability of the five drug classes, but a model combining features to predict likely tolerability has low predictive utility.

**What are the implications of our findings?:** Overall, type 2 diabetes drug tolerability cannot be accurately predicted for individual patients using routine clinical features. Prescribing decisions in type 2 diabetes should focus on drug-specific side-effect risks and differences in glycaemic and cardiometabolic benefits.

**Graphical Abstract:** 

## Introduction

The prevalence of diabetes has been steadily increasing worldwide to an estimated 800 million affected adults, with the majority of individuals being affected with type 2 diabetes (T2D), with most of these requiring drug therapies to control glycaemia levels and lower the risk of adverse complications [1, 2, 3]. Managing T2D requires multiple behavioural and pharmacological treatment strategies, including management of glycaemia, weight, cardiovascular risk factors, comorbidities, complications, and medication side-effects [4]. Metformin is currently prescribed as a first-line medication for treating T2D due to its safety profile, but as T2D progresses, a single therapy becomes insufficient [4]. T2D treatment guidelines recommend the addition of other glucose-lowering therapies – glucagon-like peptide-1 receptor agonists (GLP-1RA); dipeptidyl peptidase-4 inhibitor (DPP4i); sodium-glucose co-transporter-2 inhibitors (SGLT2i); thiazolidinediones (TZD) and sulfonylureas (SU) – when a single agent becomes insufficient [4]. Despite recent progress in T2D precision medicine [4], there is still uncertainty on the best treatment for individual patients and what metrics should be used for treatment targeting.

While recent studies have proposed targeting specific T2D medication at the individual-patient level based on differences in glycaemic benefits [5, 6], comparable data to inform targeting based on drug side-effects and/or likely tolerability is limited. All glucose-lowering agents have established specific side-effects that can frequently lead to poor tolerability and discontinuation [7, 8, 9]. Whilst the risk of each individual drug-specific side-effects can be studied directly [4, 10], an alternative is to evaluate short-term discontinuation risk as a proxy outcome measure of likely tolerability, applicable across all drug classes [5, 11]. This approach is supported by recent studies demonstrating that short-term fracture and genital infection side-effects lead to an increased risk of early discontinuation with SGLT2i [9, 12]. Although multiple other factors can lead to a perceived lack of medication efficacy or access to medication [4, 13], short-term side-effects are likely the main reason for early discontinuation.

We aimed to investigate the potential for a precision medicine approach based on routine clinical features to predict which patients are more likely to tolerate each of the five major T2D therapy classes after initial metformin, using 3-month discontinuation as a proxy outcome measure. First, we assessed differences in short-term discontinuation when stratifying by clinical and sociodemographic features and hence highlighting potential treatment heterogeneity. We then evaluated the potential for targeting specific therapy classes in individual patients by combining multiple features in a model to predict drug-specific likely tolerability.

## Methods

### Study population

Adults with T2D initiating GLP-1RA, DPP4i, SGLT2i, TZD or SU therapies of any type for the first time between 1 January 2014 and 31 October 2020 were identified in the UK population-representative Clinical Practice Research Datalink (CPRD) Aurum dataset [14] linked to Hospital Episode Statistics (HES), Office for National Statistics (ONS) death registrations and individual-level Index of Multiple Deprivation (IMD), following our previously published cohort profile [15] (see https://github.com/Exeter-Diabetes/ for all codelists and cohort preparation algorithms). We excluded individuals initiating these therapies as first-line monotherapy (as this does not align with current guidance [4]), multiple glucose-lowering therapies on the same day (as this is non-standard in the UK), missing baseline HbA_1c_ and baseline HbA_1c_ <53 mmol/mol (7%) (as the indication for treatment initiation may not be for glucose-lowering) (sFig.1).

### Outcome

The primary outcome was discontinuation within three months of initiation (a proxy for drug tolerability), with no changes to other glucose-lowering therapies during this period. Therapy discontinuation was defined as a prescription gap above 180 days. To ensure sufficient follow-up to assess this definition, patients with less than six months of follow-up data after their last prescription (due to the end of the study period, practice deregistration, or death) were excluded since discontinuation could not be confirmed. As sensitivity analyses, we also assessed discontinuation when defined at 6- and 12-months post-drug initiation.

### Candidate predictors

Candidate predictors were selected to represent routinely available sociodemographic, behavioural and clinical features [5, 6]. These comprised: current age, duration of diabetes, sex (self-reported, categorised into male, female), social deprivation (English Index of Multiple Deprivation quintile), ethnicity (self-reported, categorised into major UK groups: White, South Asian, Black, Mixed, other), smoking status (self-reported, categorised into active smoker, ex-smoker, non-smoker), baseline HbA_1c_ (closest to treatment start date; range in previous six months to +7 days), BMI, eGFR (closest to treatment start date; range in previous 2 years to +7 days), number of ever prescribed glucose-lowering therapy classes, number of current prescribed glucose-lowering therapy classes, a history of frailty (a proxy defined with a history of falls or lower limb fractures), previous prescription of blood pressure medication, previous prescription of statins (any prescription before treatment start) and history of metformin discontinuation within three months of initiation.

### Descriptive analysis

We estimated the proportion of discontinuations for each drug class stratified into subgroups defined by the predictor features. We described the proportion of individuals discontinuing each drug class by subgroups.

### Treatment selection model development

A Bayesian additive regression trees (BART) model was developed using the candidate features to predict discontinuation risk for each of the five glucose-lowering therapies. BART is a nonparametric regression modelling framework that captures predictor relationships and interactions without the need for pre-specification of exactly what form these relationships will take. This framework provides a lot of flexibility compared to more conventional methods when investigating treatment effect heterogeneity between different drug classes [16]. We used a standard BART model fitted using MCMC and the *bartMachine* package [17] (version 1.3.4.1) in R [18] (version 4.3.2). As we aimed to explore the potential for accurately predicting discontinuation, the whole cohort was used for model development and validation without resampling (or splitting the cohort into development and hold-out data). Individuals were excluded from model development if they had any missing features, as earlier work demonstrated that imputing missing values under a missing-completely-at-random assumption for a dataset of this magnitude does not markedly improve precision [19].

The discriminative power of the prediction model was assessed using the area under the receiver operating characteristic (AUROC) curves and calibration curves to ensure the predicted probability matched the observed proportions of discontinuation, overall and for each drug class. As a sensitivity analysis, separate models were also developed for discontinuation at 6- and 12-months.

Potential heterogeneous treatment effects were evaluated with conditional average treatment effects (CATE) using the concordant–discordant approach previously proposed [5, 6]. The CATE for an individual is conditional on clinical features and represents the predicted differential effects of two drug classes on the risk of discontinuation. In this approach, the cohort is split into subgroups based on predicted CATE estimates (defined by deciles), and the average CATE estimate within each subgroup is compared to estimates of average treatment effects derived from a set of alternative models fitted to each of the subgroups in turn. These latter models target the average treatment effect (ATE) within a population of individuals (rather than CATE), with desirable properties justified in the literature [20]. Alignment between both estimates provides evidence that ATEs are consistent across different inference methods within each subgroup. We used logistic regression as the primary approach for calculating ATEs within subgroups, estimating ATE as the difference in the risk of discontinuation between individuals receiving each drug class, adjusting for the full covariate set. Confidence intervals (CI) are estimated by refitting the models in bootstrapped datasets.

We followed TRIPOD+AI reporting guidance (ESM Materials) [21].

## Results

We included 135,410 people with T2D initiating glucose-lowering therapies (182,194 initiations – 16,347 GLP-1RA, 71,460 DPP4i, 50,510 SGLT2i, 5,081 TZD and 38,796 SU) (Table 1, sFig. 1). Baseline clinical features by initiated drug class are reported in Table 1. Of all treatment initiations, 26,138 (14.3%) were discontinued within the first three months since initiation. Discontinuation was highest for TZD initiations (19.6%) and lowest for DPP4i initiations (12.8%) (14.4% GLP-1RA, 14.9% SGLT2i, 15.7% SU). Discontinuation was higher at 6-months (range 17.2%-26.2%) and 12-months (23.2%-34.8%), but the relative proportions discontinuing each of the five drug classes was similar (Figure 1, sTable 1).

**Table 1:** Demographic and clinical features of patients initiating the five glucose-lowering drug classes. All features were evaluated as potential predictors of discontinuation in model development. Data are mean [SD] and number (%). *Closest values to treatment start in the previous 6-months. A recorded baseline HbA_1c_ was a study inclusion criteria.

|  | GLP-1RA<br>(n=16,347) | DPP4i<br>(n=71,460) | SGLT2i<br>(n=50,510) | TZD<br>(n=5,081) | SU<br>(n=38,796) |
| --- | --- | --- | --- | --- | --- |
| Current age, years | 58.6 [11.0] | 64.2 [12.6] | 58.7 [10.5] | 61.4 [12.1] | 61.4 [12.6] |
| Duration of diabetes, years | 9.5 [5.8] | 8.6 [5.8] | 8.9 [5.6] | 8.1 [4.9] | 6.2 [4.6] |
| Sex |  |  |  |  |  |
| Male | 8,650 (52.9) | 42,218 (59.1) | 30,303 (60.0) | 3,117 (61.3) | 22,889 (59.0) |
| Female | 7,697 (47.1) | 29,242 (40.9) | 20,207 (40.0) | 1,964 (38.7) | 15,907 (41.0) |
| Smoking status |  |  |  |  |  |
| Active | 2,575 (15.8) | 10,946 (15.3) | 8,293 (16.4) | 836 (16.5) | 6,781 (17.5) |
| Ex-smoker | 9,610 (58.8) | 40,915 (57.3) | 28,458 (56.3) | 2,771 (54.5) | 20,987 (54.1) |
| Non-smoker | 3,470 (21.2) | 16,227 (22.7) | 11,760 (23.3) | 1,235 (24.3) | 9,251 (23.8) |
| Missing | 692 (4.2) | 3,372 (4.7) | 1,999 (4.0) | 239 (4.7) | 1,777 (4.6) |
| Ethnicity |  |  |  |  |  |
| White | 14,305 (87.5) | 57,903 (81.0) | 40,802 (80.8) | 4,068 (80.1) | 31,299 (80.7) |
| South Asian | 1,062 (6.5) | 7,647 (10.7) | 5,749 (11.4) | 625 (12.3) | 4,212 (10.9) |
| Black | 519 (3.2) | 3,054 (4.3) | 1,693 (3.4) | 176 (3.5) | 1,776 (4.6) |
| Other | 114 (0.7) | 833 (1.2) | 594 (1.2) | 54 (1.1) | 484 (1.2) |
| Mixed | 107 (0.7) | 579 (0.8) | 418 (0.8) | 43 (0.8) | 304 (0.8) |
| Missing | 240 (1.5) | 1,444 (2.0) | 1,254 (2.5) | 115 (2.3) | 721 (1.9) |
| Index of multiple deprivation |  |  |  |  |  |
| 1 (least deprived) | 2,796 (17.1) | 12,476 (17.5) | 8,955 (17.7) | 921 (18.1) | 6,380 (16.4) |
| 2 | 2,853 (17.5) | 13,092 (18.3) | 9,091 (18.0) | 923 (18.2) | 6,998 (18.0) |
| 3 | 3,097 (18.9) | 13,678 (19.1) | 9,612 (19.0) | 1,015 (20.0) | 7,318 (18.9) |
| 4 | 3,484 (21.3) | 15,482 (21.7) | 10,946 (21.7) | 1,046 (20.6) | 8,652 (22.3) |
| 5 (most deprived) | 4,117 (25.2) | 16,732 (23.4) | 11,906 (23.6) | 1,176 (23.1) | 9,448 (24.4) |
| Biomarkers |  |  |  |  |  |
| BMI, kg/m<sup>2</sup> | 37.2 [7.0] | 32.0 [6.6] | 34.1 [6.8] | 31.8 [6.4] | 32.1 [6.6] |
| Missing | 432 (2.6) | 3,372 (4.7) | 1,707 (3.4) | 185 (3.6) | 2,256 (5.8) |
| HbA<sub>1c</sub>, mmol/mol* | 79.5 [16.0] | 73.7 [15.5] | 77.7 [16.1] | 76.7 [15.7] | 79.8 [19.8] |
| Missing* | - | - | - | - | - |
| eGFR, ml/min per 1.73m<sup>2</sup> | 90.3 [20.6] | 84.1 [22.6] | 94.4 [15.3] | 88.2 [21.0] | 88.8 [20.8] |
| Missing | 20 (0.1) | 112 (0.2) | 82 (0.2) | 5 (0.1) | 90 (0.2) |
| History of statins | 14,292 (87.4) | 62,035 (86.8) | 43,395 (85.9) | 4,444 (87.5) | 32,069 (82.7) |
| History of blood medication | 1,780 (10.9) | 10,809 (15.1) | 6,999 (13.9) | 755 (14.9) | 5,463 (14.1) |
| History of frailty | 3,569 (21.8) | 16,201 (22.7) | 9,691 (19.2) | 961 (18.9) | 7,928 (20.4) |
| History of metformin discontinuation with 3-months | 2,267 (13.9) | 9,274 (13.0) | 6,838 (13.5) | 746 (14.7) | 4,585 (11.8) |
| Number of other<br>current glucose-lowering drugs |  |  |  |  |  |
| 0 | 706 (4.3) | 6,619 (9.3) | 2,028 (4.0) | 274 (5.4) | 4,608 (11.9) |
| 1 | 5,490 (33.6) | 43,283 (60.6) | 20,325 (40.2) | 1,897 (37.3) | 26,725 (68.9) |
| 2+ | 10,151 (62.1) | 21,558 (30.2) | 28,157 (55.7) | 2,910 (57.3) | 7,463 (19.2) |
| Number of glucose-lowering drug<br>classes ever<br>prescribed<br>(including therapy<br>initiated) |  |  |  |  |  |
| 2 | 1,721 (10.5) | 32,882 (46.0) | 11,717 (23.2) | 990 (19.5) | 26,154 (67.4) |
| 3 | 3,703 (22.7) | 27,943 (39.1) | 14,577 (28.9) | 1,995 (39.3) | 9,447 (24.4) |
| 4+ | 10,923 (66.8) | 10,635 (14.9) | 24,216 (47.9) | 2,096 (41.3) | 3,195 (8.2) |

**Fig. 1:**
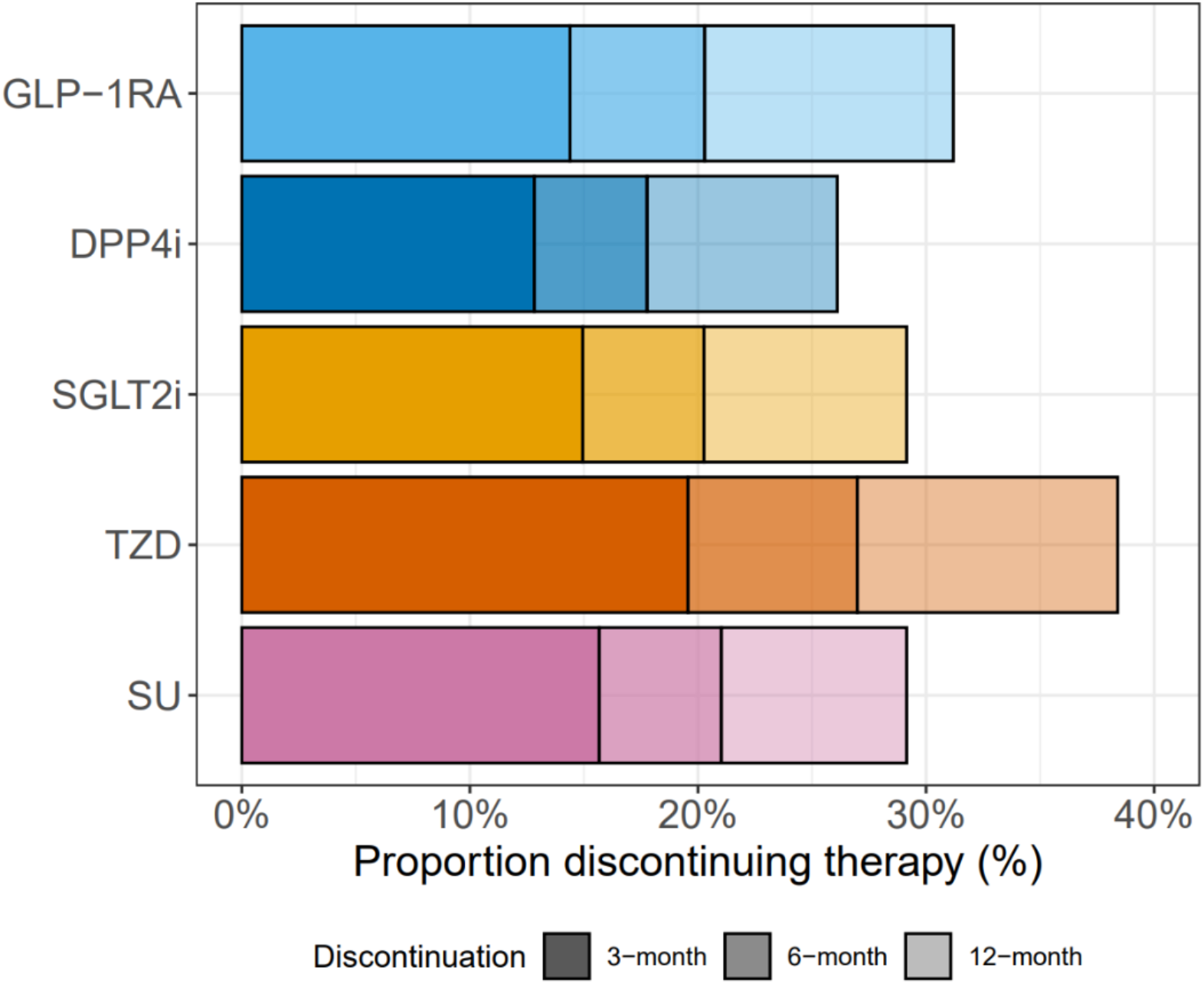
Proportion of individuals with type 2 diabetes discontinuing the five drug classes, at 3-months (primary outcome), 6-months, and 12-months. See sTable 1 for a breakdown of patient numbers for each therapy at each time point.

### 3-month discontinuation varied across the five therapy classes by age and routine clinical features

Figure 2 shows the proportions of patients discontinuing therapy within 3-months stratified by sociodemographic and clinical features. When assessing sociodemographic features, there was some evidence that age at treatment initiation was associated with differences in discontinuation by drug, with higher discontinuation of GLP-1RA and SGLT2i in older adults, which was not observed for the other therapies. Discontinuation was higher in females and individuals of South Asian and Black ethnicity compared to White ethnicity, with no evidence of variation across the five therapy classes. Discontinuation was consistent by deprivation and smoking status. In terms of clinical features, we found higher discontinuation of GLP-1RA and SGLT2i in patients with longer duration of diabetes, lower discontinuation of SGLT2i, TZD and SU as the number of other current glucose-lowering therapies increased and higher discontinuation of SGLT2i, DPP4i and TZD as the number of previously prescribed therapies increased. Higher discontinuation was seen for patients not taking statins or blood pressure medication and those who have discontinued metformin in the past (similar across the five drug classes). When assessing laboratory measures, there was evidence of higher discontinuation at higher levels of HbA_1c_ for DPP4i and TZD and higher discontinuation at lower levels of BMI for GLP-1RA, TZD and SU (Fig. 2, sTable 2). As a sensitivity analysis, the 6- and 12-months discontinuation patterns across all features were similar to those seen for 3-month discontinuation (sFig. 2).

**Fig. 2:**
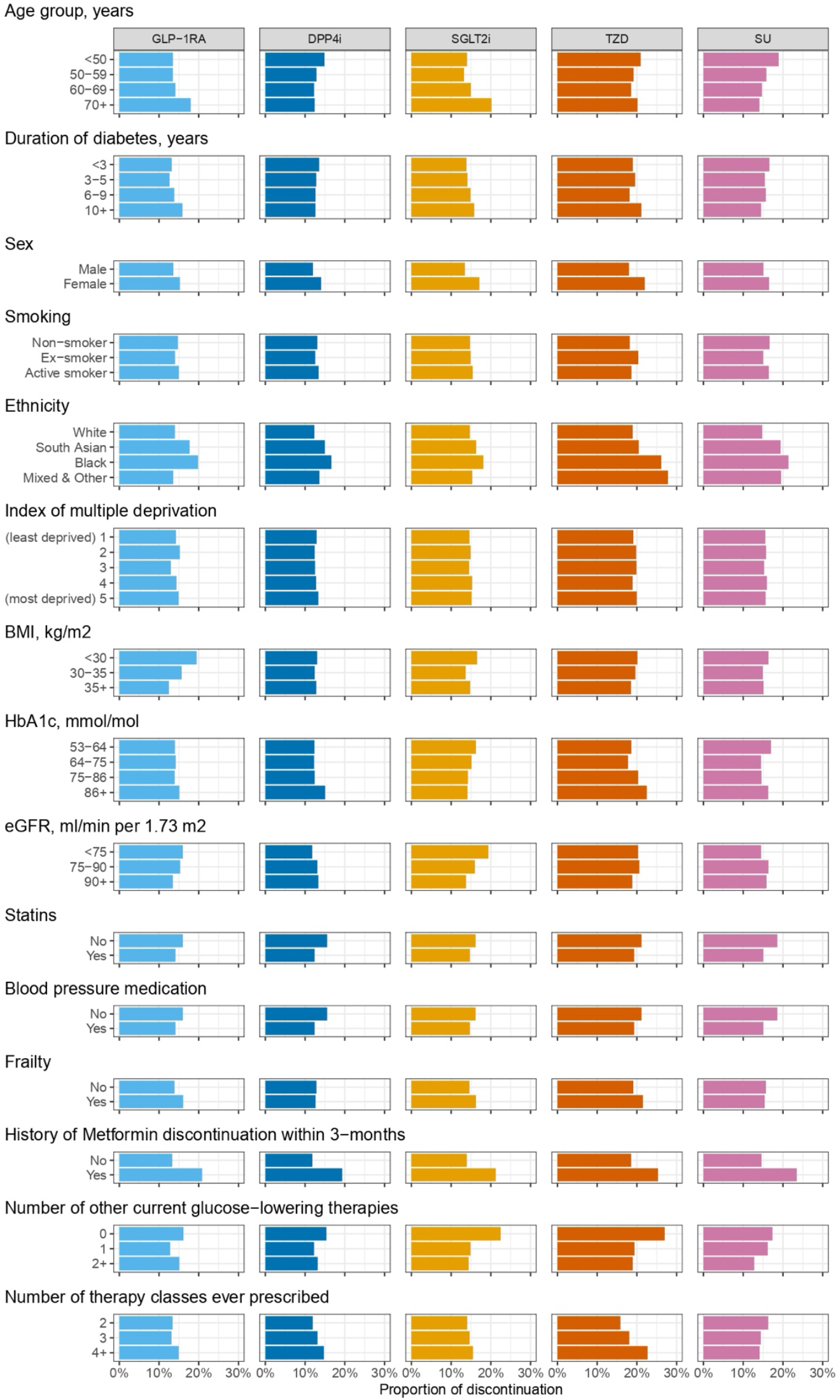
Proportion of individuals with T2D discontinuing the five drug classes at 3-months, stratified by sociodemographic and clinical features. A breakdown of patient numbers can be found in Table 1 and sTable 2.

### Discontinuation model development and validation

The discontinuation model was developed using the whole cohort (n=162,779) with valid 3-month discontinuation outcome data and complete predictor features. As sensitivity analysis, model development was repeated for 6-months (n=157,327) and 12-months (n=142,610) discontinuation outcomes.

The BART model converged quickly, and we ran 25,000 iterations, with the first 15,000 discarded as burn-in (trace plots are available on request). As would be anticipated for a within-sample evaluation, the calibration of the tolerability model was good, with the predicted discontinuation matching the observed discontinuation overall and per-therapy (Fig. 3) and ranging from 8.8% to 23.5% across deciles of predicted discontinuation probabilities. Calibration was similarly good when predicting discrimination of individual therapy classes and for the 6- and 12-months discontinuation models (sFig. 3). However, model discrimination was weak. The 3-month discontinuation outcome model achieved an overall AUROC value of 0.61 (95%CI 0.61;0.62) (Fig. 3), suggesting low utility to predict discontinuation accurately.

**Fig. 3:**
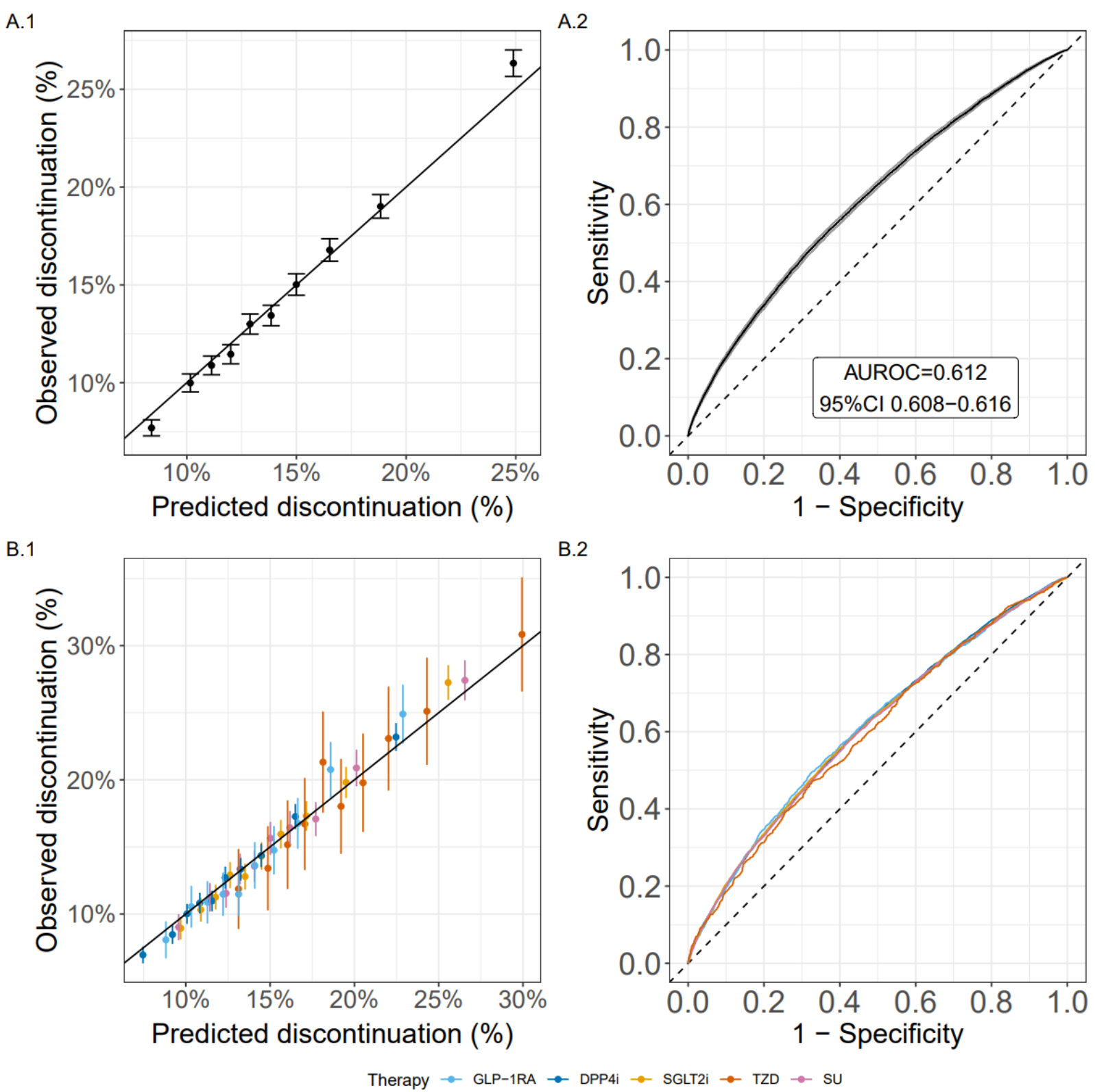
Calibration (1) and discrimination (2) plots for the model predicting discontinuation at 3-months, for overall (A) and per therapy (B) discontinuation. (1) Calibration of predicted and observed discontinuation for groups defined by decile of predicted discontinuation risk. Black line represents perfect calibration. (2) Receiver operating characteristic curve showing the discrimination between discontinuation and non-discontinuation. AUROC = area under the receiver operating characteristic curve.

Discrimination was similarly low when assessing individual therapy classes (AUROC range 0.60–0.61, Fig.3, sTable 3). Weak discrimination was also found for the 6-months (AUROC 0.61 [95%CI 0.61;0.62]) and 12-months (AUROC 0.61 [95%CI 0.61;0.62]) outcome models (sFig. 4, sTable 3).

The mean CATE between therapies combinations ranged from 0.4% – 5.6 and the model demonstrated some heterogeneity in the predicted CATE estimates (sFig. 5). However, calibration by decile of model-predicted CATE estimates often did not align with estimates of ATE (sFig. 6). A similar finding was seen for 6- and 12-months discontinuation models (sFig. 7-8).

## Discussion

In this large population-based study, we found that, although sociodemographic and clinical features are associated with differences in short-term discontinuation among new users of five major T2D drug classes (SGLT2i, GLP-1RA, DPP4i, TZD and SU), a model combining clinical features to predict differences in discontinuation risk by therapy for individual patients had low predictive ability (AUROC 0.61) and showed poor calibration of heterogeneous treatment effects. This finding suggests a limited potential for a precision medicine approach to T2D treatment based on short-term drug tolerability. The predictors with the largest differences in discontinuation across the five drug classes were age and diabetes duration, with higher discontinuation seen amongst older patients and, for SGLT2i and GLP-1RA, longer diabetes duration. Other features generally showed modest differences in short-term discontinuation that were broadly consistent by drug class, including higher discontinuation for patients who had previously discontinued metformin, females and patients of South-Asian/Black ethnicities, as well as lower discontinuation for patients currently taking statins and blood pressure medication. However, we showed that combining these differences into a prediction model was, even within sample, insufficient to predict discontinuation at the individual patient level.

Although previous studies have examined features associated with discontinuation for individual therapy classes [7, 22, 23, 24, 25], our study is the first to describe clinical features associated with discontinuation considering all five therapy classes and the first to evaluate the utility of predicting treatment discontinuation at the individual patient level. Estimates of discontinuation at 12-months are similar to previous UK studies [7, 8, 9, 26, 27]. This is likely to reflect the healthcare system in the UK, as shown by differences with data from other countries, such as higher discontinuation in the CHOICE study (six European countries) [22] and lower discontinuation in a Danish cohort [24]. Additionally, differences in discontinuation levels between studies could be attributed to the low numbers of patients by drug class and differences in features (more males, lower duration of diabetes and lower HbA_1c_) [24]. Previous studies examining factors associated with discontinuation have, in keeping with our analysis, identified higher discontinuation at older ages and lower BMI [23], and higher discontinuation for patients initiating SGLT2i with lower eGFR and frailty [9].

Our study has limitations. Although our outcome definition was informed by previous studies [8, 27], our dataset from routine clinical care did not allow us to capture the true underlying reason for discontinuation. Hence, we cannot be sure whether it represents poor tolerability or other factors. An alternative definition could have included a dose reduction (which may relate to a lack of tolerability); however, robust dosage data was unavailable in our dataset. Similarly, a diagnosis of adverse events leading to discontinuation could be used as part of the outcome definition; however, there are difficulties associated with differentiating between treatment-limiting and non-limiting adverse events in primary care data [28]. However, sensitivity analysis suggests our findings on overall discontinuation levels are consistent with previous UK studies [7, 8, 9], supporting our choice. The requirement for a gap in prescriptions above 180 days means we can reasonably ensure the treatment has been discontinued for a sustained period. However, it does not fully distinguish between temporary and permanent discontinuation [29]. Reassuringly, all our analyses were consistent when assessing discontinuation at 6- and 12-months, supporting the generalisability of our outcome definition. This study deliberately only looked at baseline features as we aimed to establish who might discontinue when initiating a new drug to potentially inform baseline treatment decisions, and we did not intend to look at what might affect discontinuation after treatment has started, unlike previous work [30]. This study contains lower numbers of TZD initiations versus other therapies, but compared to other studies, this study includes higher numbers of therapy initiations for all therapies [7, 8]. Finally, as our analysis was performed using UK data, findings may not be generalisable to other countries and health systems.

This is the first study to explore differential discontinuation between all five major glucose-lowering therapies using routine clinical features, providing notable evidence of higher discontinuation for older adults starting GLP-1RA and SGLT2i. Importantly, we showed that routine clinical features do not provide enough clinical utility alone to inform treatment targeting by predicting patients at high risk of short-term discontinuation of glucose-lowering therapy. The model developed in this study could be deployed in conjunction with other treatment selection tools [5, 6, 11] to provide further differentiation in situations of equipoise. Despite this, our analysis does not undermine the importance of robust studies to understand susceptibility to drug-specific side-effects, which could include genetics and multi-omics [31, 32]. Considering treatment benefits, recent work has identified practical applications of routine clinical features to target T2D medication based on glucose-lowering, with robust heterogenous treatment effects for all five drug classes, providing clear evidence to guide choice of therapy for individual patients [5, 6, 11, 33, 34, 35, 36].

## Conclusion

Our study demonstrates that short-term discontinuation varies modestly across the five major T2D drug classes after metformin by certain clinical features, including age and diabetes duration. Despite this variation, our analysis suggests a limited scope to predict the likelihood of short-term tolerability by drug class for individual patients. This indicates that T2D clinical prescribing decisions should focus on drug-specific glycaemic and cardiometabolic benefits, alongside specific side-effect risks, instead of potential tolerability.

## Supporting information

sFig

## Data Availability

The UK routine clinical data analysed during the current study are available in the CPRD repository (CPRD; https://cprd.com/research-applications), but restrictions apply to the availability of these data, which were used under license for the current study, and so are not publicly available. For re-using these data, an application must be made directly to CPRD. All R code used for the analysis is provided at https://github.com/Exeter-Diabetes/CPRD-Pedro-T2DDiscontinuation.

## Acknowledgements

For the purpose of open access, the author has applied a CC BY public copyright license to any Author Accepted Manuscript version arising from this submission. This article is based in part on data from the CPRD obtained under license from the UK Medicines and Healthcare products Regulatory Agency. CPRD data are provided by patients and collected by the UK National Health Service (NHS) as part of their care and support. Approval for CPRD data access and the study protocol was granted by the CPRD Independent Scientific Advisory Committee (eRAP protocol number: 22_002000). The authors acknowledge contributions from the wider MASTERMIND consortium who supported this work (see ESM for a list of MASTERMIND consortium members).

## Funding and Assistance

The authors acknowledge support from the Medical Research Council (UK) (MR/N00633X/1), and the National Institute for Health and Care Research Exeter Biomedical Research Centre. JMD is supported by a Wellcome Trust Early Career award (227070/Z/23/Z). ATH and BMS are supported by the NIHR Exeter Clinical Research Facility. The views expressed are those of the authors and not necessarily those of the NHS, the NIHR or the Department of Health.

## Author Contributions

PC, JMD, BMS, ATH conceived and designed the study. PC analysed the data and developed the code. KGY and RH helped with curating the CPRD dataset. All authors contributed to the writing of the article, provided support for the analysis and interpretation of results, critically revised the article and approved the final article.

