## Supplementary material for "Evaluating prediction of short-term tolerability of five type 2 diabetes drug classes using routine clinical features: UK population-based study": sFig

### Supplementary Materials:

#### Tables:

**sTable 1: Therapy discontinuation at 3-months (primary outcome), 6-months, and 12-months.** N's represent the number of drug initiations included in the analysis of 3-month discontinuation. Breakdown of patient numbers for each therapy at each time point. Data are number (%).

| Therapy | GLP-1RA (n=16,347) | DPP4i (n=71,460) | SGLT2i (n=50,510) | TZD (n=5,081) | SU (n=38,796) |
|---|---|---|---|---|---|
| 3-month discontinuation | | | | | |
| No | 13,995 (85.6) | 62,296 (87.2) | 42,961 (85.1) | 4,087 (80.4) | 32,717 (84.3) |
| Yes | 2,352 (14.4) | 9,164 (12.8) | 7,549 (14.9) | 994 (19.6) | 6,079 (15.7) |
| 6-month discontinuation | | | | | |
| No | 12,464 (76.2) | 56,958 (79.7) | 38,761 (76.7) | 3,604 (70.9) | 29,694 (76.5) |
| Yes | 3,173 (19.4) | 12,306 (17.2) | 9,850 (19.5) | 1,332 (26.2) | 7,904 (20.4) |
| Missing | 710 (4.3) | 2,196 (3.1) | 1,899 (3.8) | 145 (2.9) | 1,198 (3.1) |
| 12-month discontinuation | | | | | |
| No | 9,387 (57.4) | 46,957 (65.7) | 30,467 (60.3) | 2,836 (55.8) | 24,770 (63.8) |
| Yes | 4,256 (26.0) | 16,589 (23.2) | 12,534 (24.8) | 1,768 (34.8) | 10,190 (26.3) |
| Missing | 2,704 (16.5) | 7,914 (11.1) | 7,509 (14.9) | 477 (9.4) | 3,836 (9.9) |

**sTable 2: Breakdown of patient numbers across subgroups of candidate predictors in Figure 2.** Data are number (%).

| Therapy | GLP-1RA (n=16,347) | DPP4i (n=71,460) | SGLT2i (n=50,510) | TZD (n=5,081) | SU (n=38,796) |
|---|---|---|---|---|---|
| Current age, years | | | | | |
| <50 | 3,361 (20.6) | 9,906 (13.9) | 9,979 (19.8) | 883 (17.4) | 7,387 (19.0) |
| 50-59 | 5,555 (34.0) | 17,103 (23.9) | 17,483 (34.6) | 1,480 (29.1) | 10,525 (27.1) |
| 60-69 | 4,879 (29.8) | 19,792 (27.7) | 15,739 (31.2) | 1,439 (28.3) | 10,687 (27.5) |
| 70+ | 2,552 (15.6) | 24,659 (34.5) | 7,309 (14.5) | 1,279 (25.2) | 10,197 (26.3) |
| Duration of diabetes, years | | | | | |
| <3 | 2,014 (12.3) | 12,317 (17.2) | 7,347 (14.5) | 753 (14.8) | 10,908 (28.1) |
| 3-5 | 3,237 (19.8) | 15,558 (21.8) | 10,585 (21.0) | 1,176 (23.1) | 10,583 (27.3) |
| 6-9 | 4,361 (26.7) | 18,210 (25.5) | 13,322 (26.4) | 1,535 (30.2) | 9,765 (25.2) |
| 10+ | 6,735 (41.2) | 25,375 (35.5) | 19,256 (38.1) | 1,617 (31.8) | 7,540 (19.4) |
| Biomarkers | | | | | |
| BMI, kg/$m^2$ | | | | | |
| <30 | 1,861 (11.4) | 28,533 (39.9) | 13,994 (27.7) | 2,138 (42.1) | 15,139 (39.0) |
| 30-35 | 4,837 (29.6) | 20,967 (29.3) | 16,033 (31.7) | 1,443 (28.4) | 11,233 (29.0) |
| 35+ | 9,217 (56.4) | 18,588 (26.0) | 18,776 (37.2) | 1,315 (25.9) | 10,168 (26.2) |
| Missing | 432 (2.6) | 3,372 (4.7) | 1,707 (3.4) | 185 (3.6) | 2,256 (5.8) |
| $HbA_{1c}$, mmol/mol* | | | | | |
| 53-64 | 2,516 (15.4) | 21,314 (29.8) | 10,009 (19.8) | 997 (19.6) | 8,536 (22.0) |
| 64-75 | 4,782 (29.3) | 23,851 (33.4) | 15,602 (30.9) | 1,777 (35.0) | 11,122 (28.7) |
| 75-86 | 3,996 (24.4) | 12,831 (18.0) | 11,209 (22.2) | 1,092 (21.5) | 7,173 (18.5) |
| 86+ | 5,053 (30.9) | 13,464 (18.8) | 13,690 (27.1) | 1,215 (23.9) | 11,965 (30.8) |
| eGFR, ml/min per 1.73$m^2$ | | | | | |
| <75 | 3,562 (21.8) | 21,751 (30.4) | 6,175 (12.2) | 1,206 (23.7) | 9,083 (23.4) |
| 75-90 | 3,067 (18.8) | 15,116 (21.2) | 10,953 (21.7) | 1,028 (20.2) | 7,834 (20.2) |
| 90+ | 9,698 (59.3) | 34,481 (48.3) | 33,300 (65.9) | 2,842 (55.9) | 21,789 (56.2) |
| Missing | 20 (0.1) | 112 (0.2) | 82 (0.2) | 5 (0.1) | 90 (0.2) |

**sTable 3: Discrimination values (AUC) for discontinuation BART models at 3-months, 6-months and 12-months.** Models were fitted to all therapies simultaneously.

| Model | 3-month discontinuation | 6-month discontinuation | 12-month discontinuation |
|---|---|---|---|
| Overall | 0.612 (0.608; 0.616) | 0.611 (0.608; 0.615) | 0.614 (0.610; 0.617) |
| GLP-1RA | 0.610 (0.596; 0.623) | 0.594 (0.583; 0.606) | 0.585 (0.574; 0.596) |
| DPP4i | 0.609 (0.602; 0.616) | 0.607 (0.601; 0.613) | 0.613 (0.607; 0.618) |
| SGLT2i | 0.608 (0.600; 0.615) | 0.611 (0.605; 0.618) | 0.614 (0.608; 0.620) |
| TZD | 0.596 (0.575; 0.617) | 0.612 (0.593; 0.631) | 0.607 (0.590; 0.625) |
| SU | 0.605 (0.597; 0.614) | 0.603 (0.595; 0.610) | 0.605 (0.598; 0.612) |

### Figures:

**sFig. 1: CPRD patient flow and inclusion criteria for the analysis cohorts.** Baseline $HbA_{1c}$ is defined as the closest $HbA_{1c}$ to drug initiation in the previous 6-months. Other biomarkers were defined as the closest measure to drug initiation in the previous 2 years.

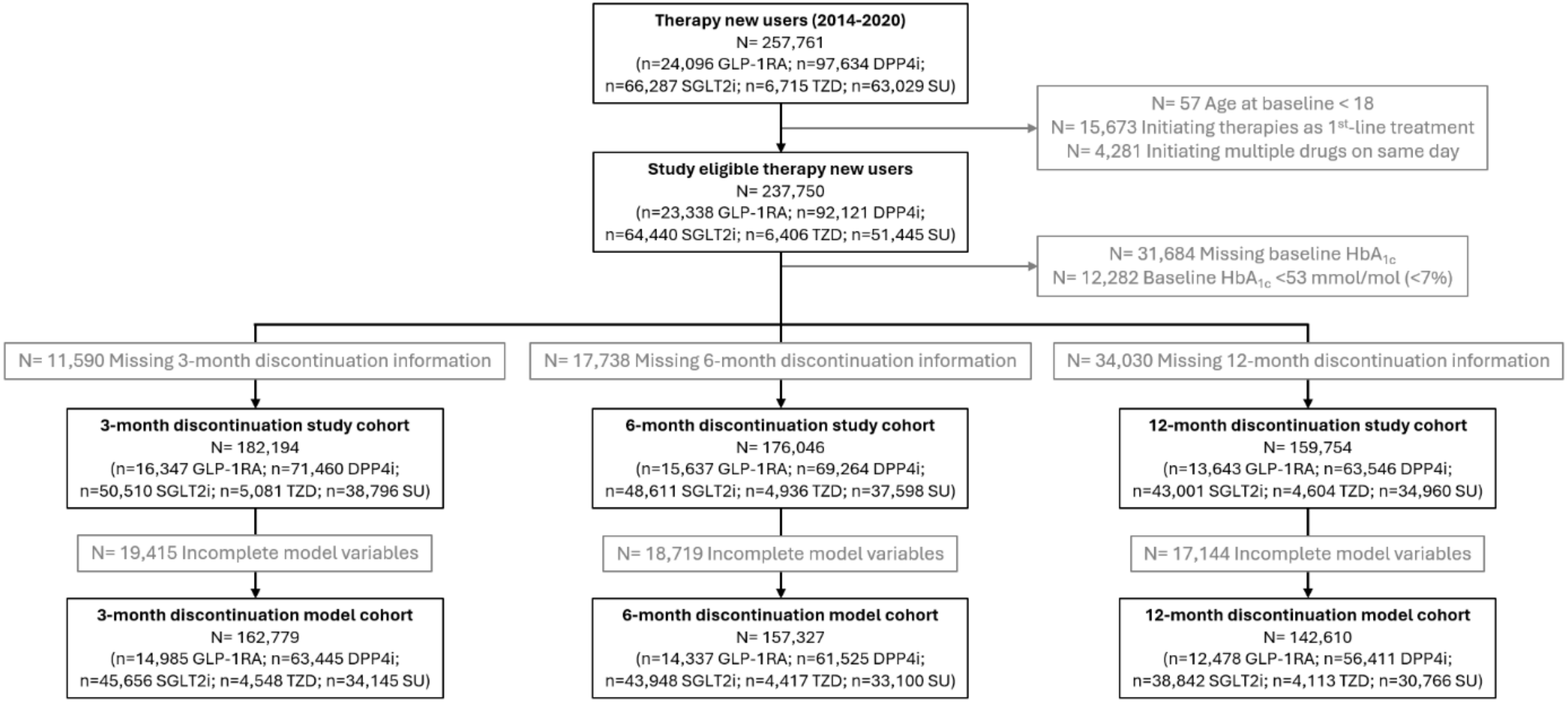

**sFig. 2: Proportion of individuals with T2D discontinuing the five drug classes at 3-, 6- and 12-months, stratified by sociodemographic and clinical features.**

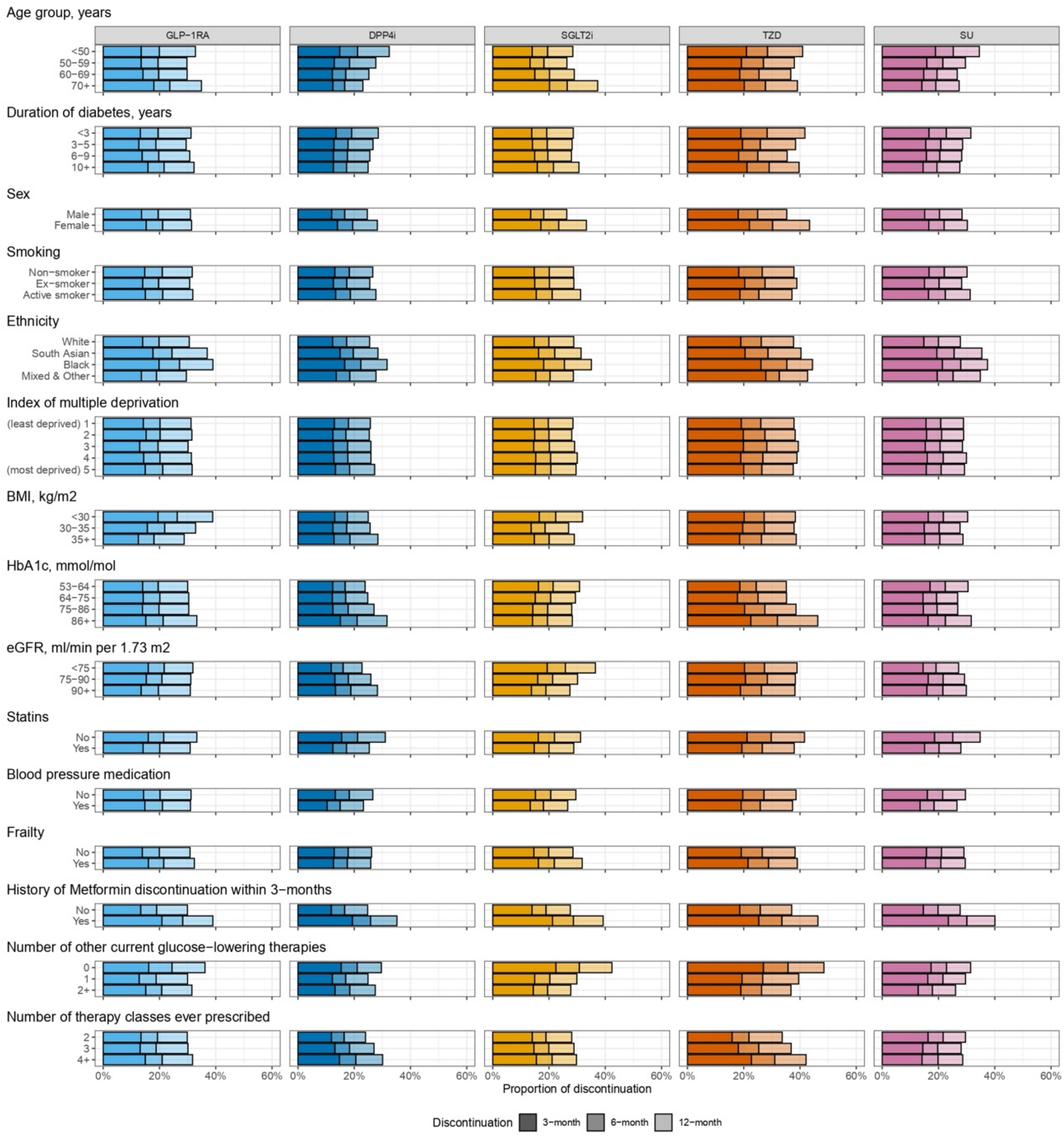

**sFig. 3: Calibration plots for discontinuation BART models at 3-months (A), 6-months (B) and 12-months (C), for overall (1) and per therapy (2) discontinuation.** (A) discontinuation at 3-months. (B) discontinuation at 6-months. (C) discontinuation at 12-months. (1) overall discontinuation. (2) per therapy discontinuation. Calibration plot shows the predicted and observed discontinuation for groups defined by decile of predicted discontinuation risk. Black line represents perfect calibration.

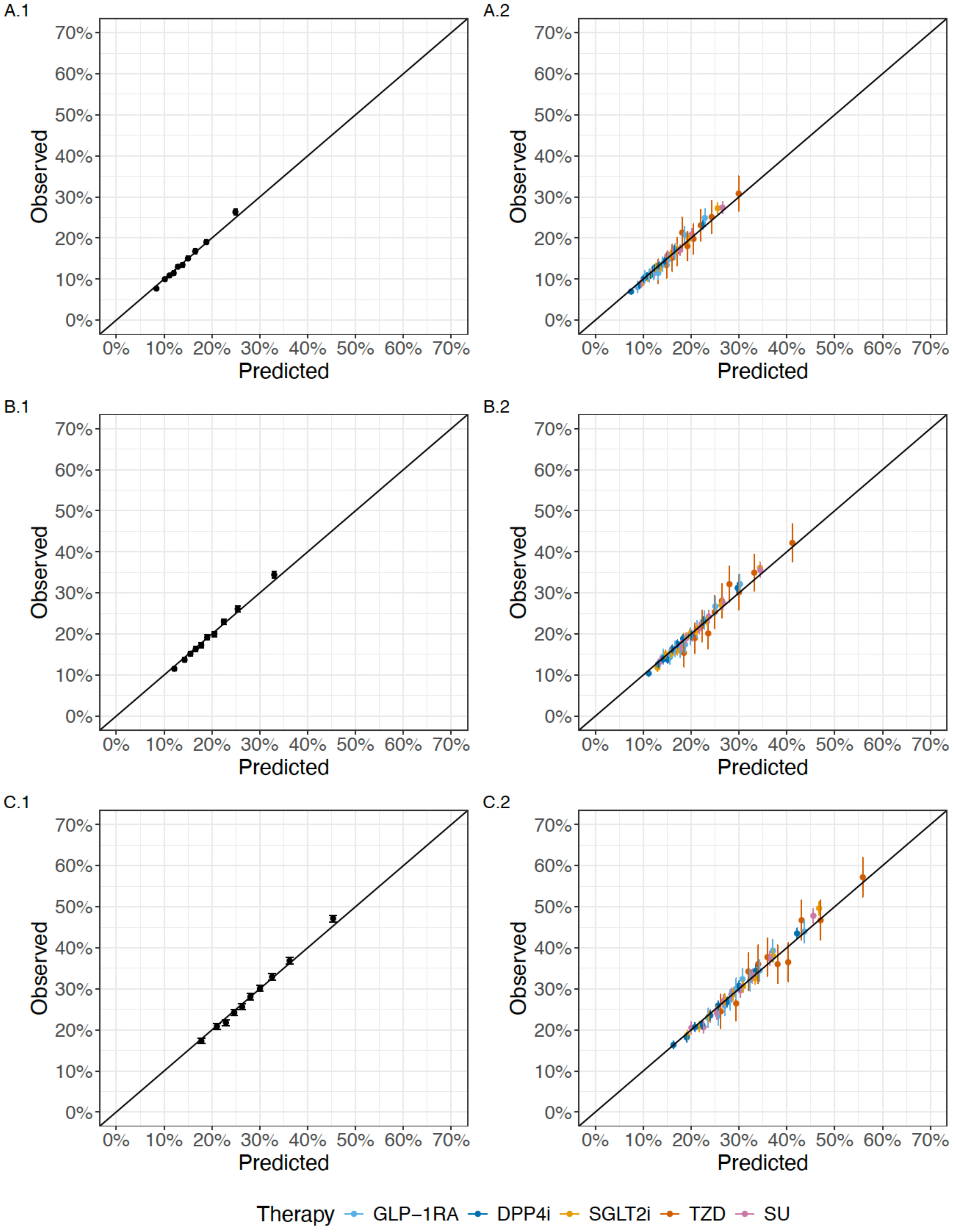

**sFig. 4: Discrimination plots for discontinuation BART models at 3-months (A), 6-months (B) and 12-months (C), for overall (1) and per therapy (2) discontinuation.** (A) discontinuation at 3-months. (B) discontinuation at 6-months. (C) discontinuation at 12-months. (1) overall discontinuation. (2) per therapy discontinuation. AUROC = aura under the receiver operating characteristics curve. A breakdown of AUC values can be found in sTable 3.

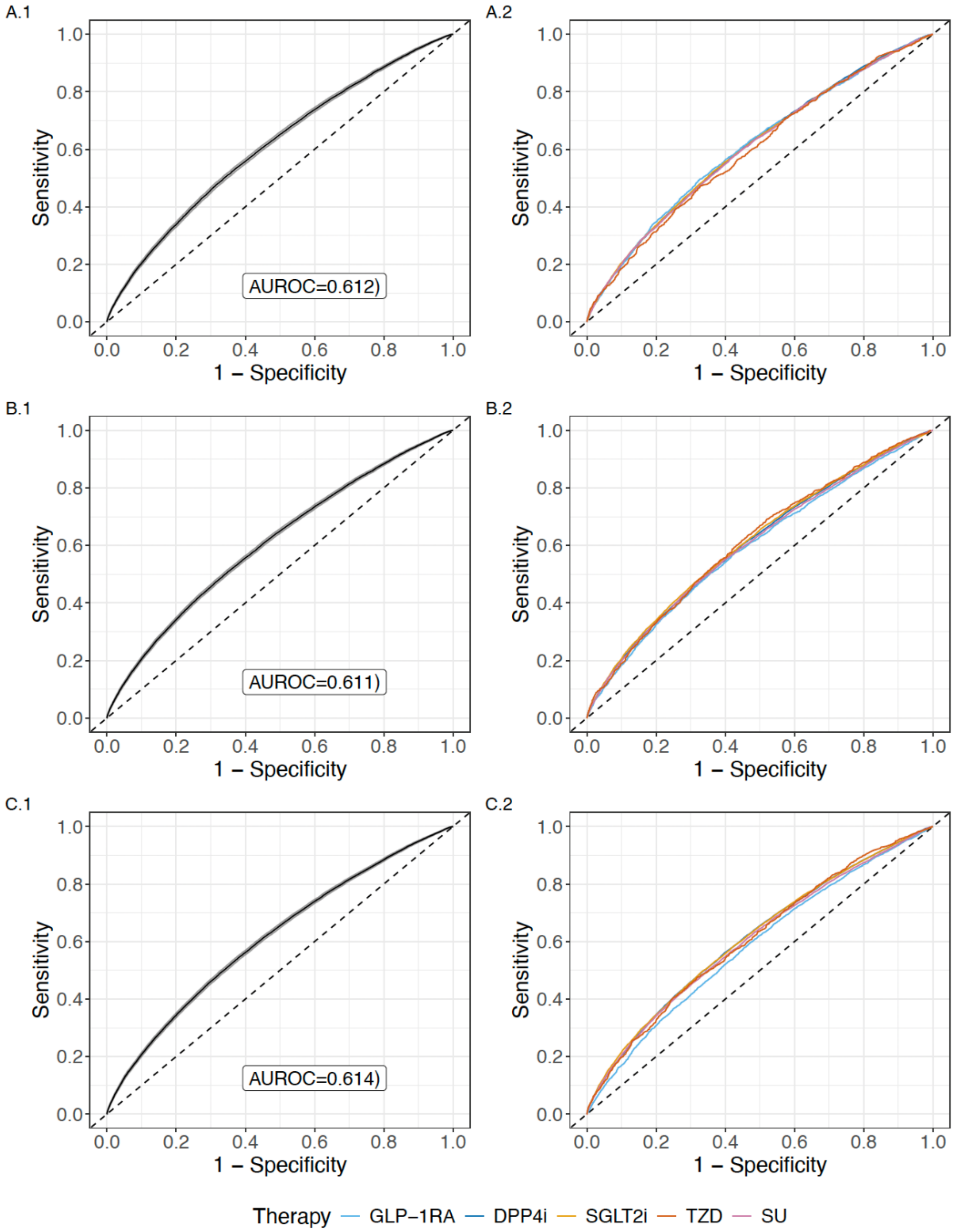

**sFig. 5: Distribution of predicted CATE effects for drug pairs.** Negative values reflect a predicted HbA1c treatment benefit on drug A (left side) and positive values reflect a predicted treatment benefit on drug B (right side).

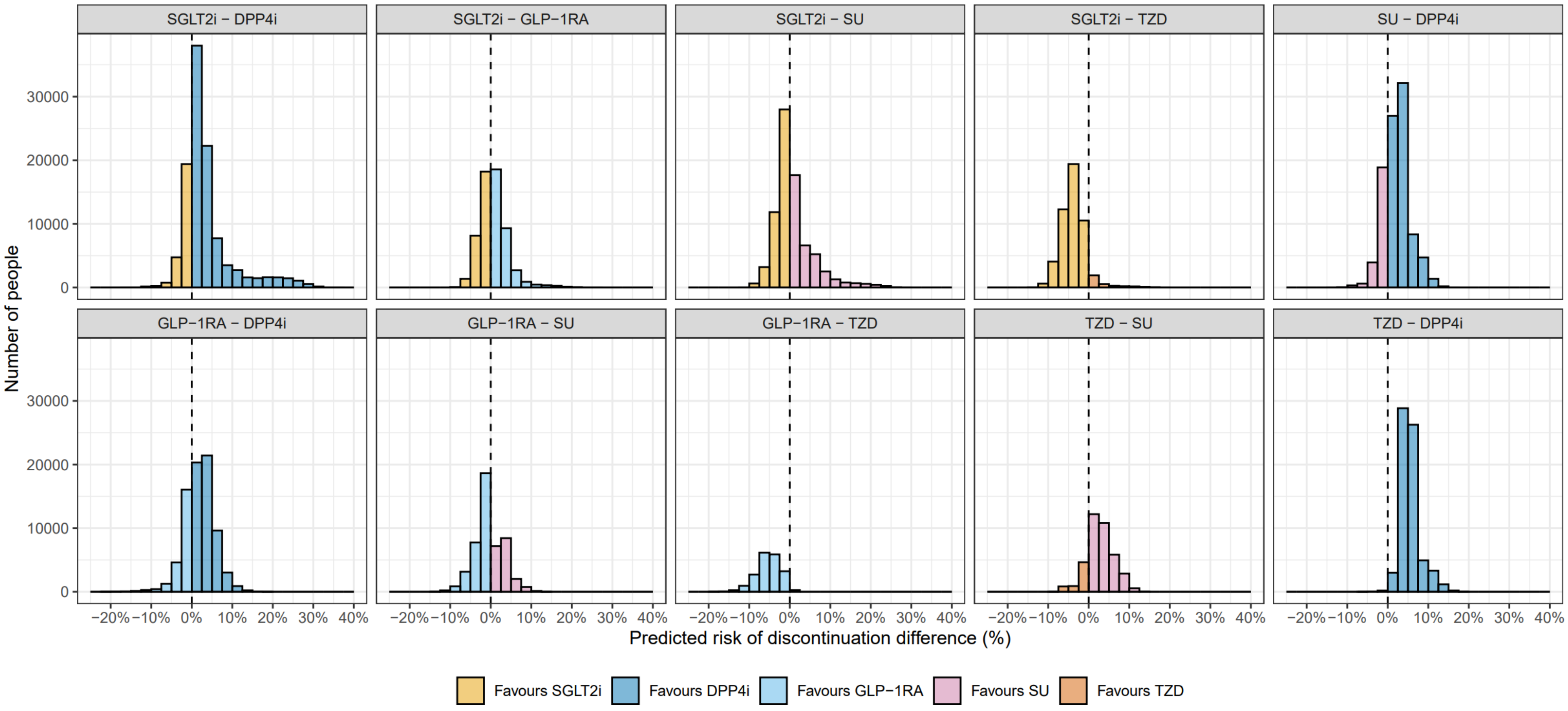

**sFig. 6: Calibration of predicted heterogeneous treatment effects across drug class pairs for 3-month discontinuation.** Red line represents perfect calibration. Point estimates represent predicted and observed differences in 3-month discontinuation. Error bars represent 95% Cis calculated through bootstrapping.

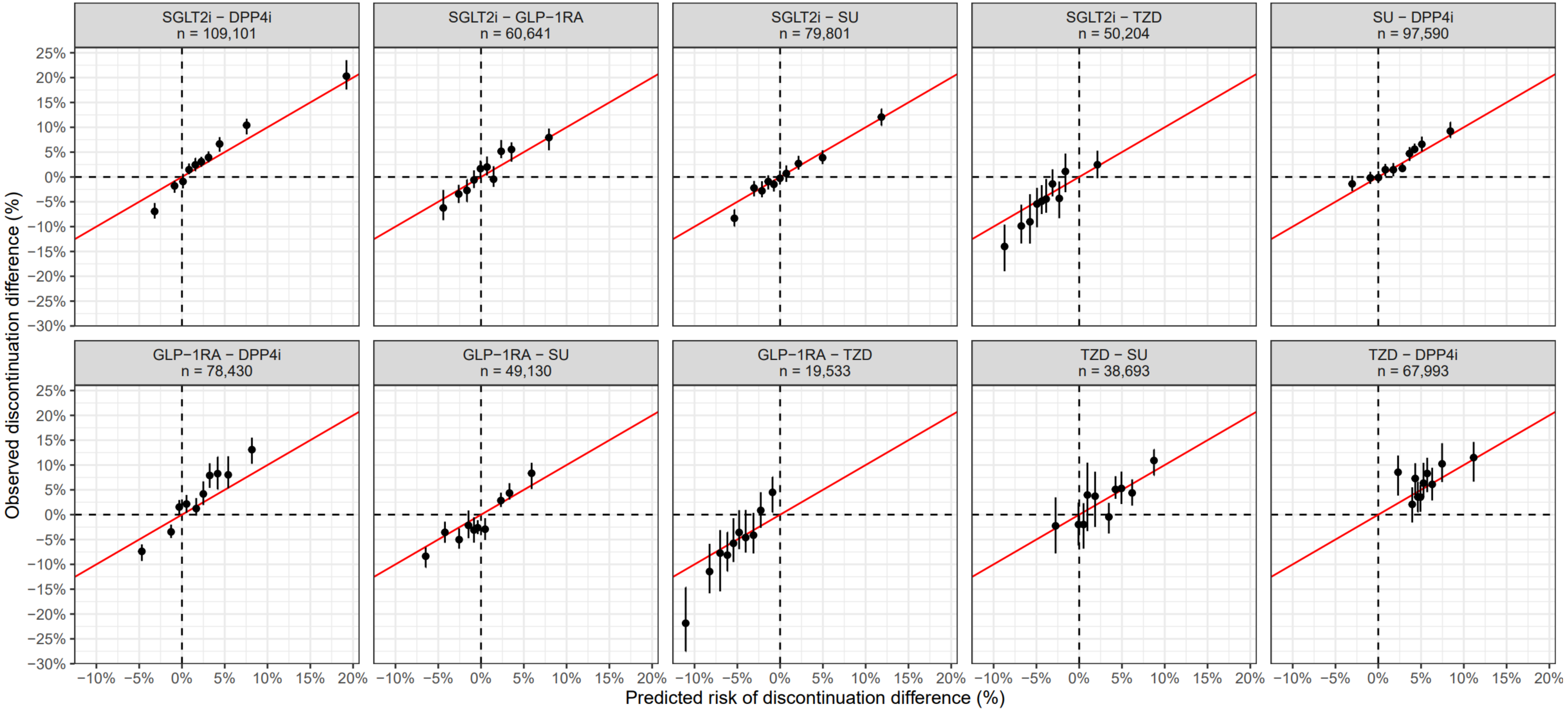

**sFig. 7: Calibration of predicted heterogeneous treatment effects across drug class pairs for 6-month discontinuation.** Red line represents perfect calibration. Point estimates represent predicted and observed differences in 6-month discontinuation. Error bars represent 95% Cis calculated through bootstrapping.

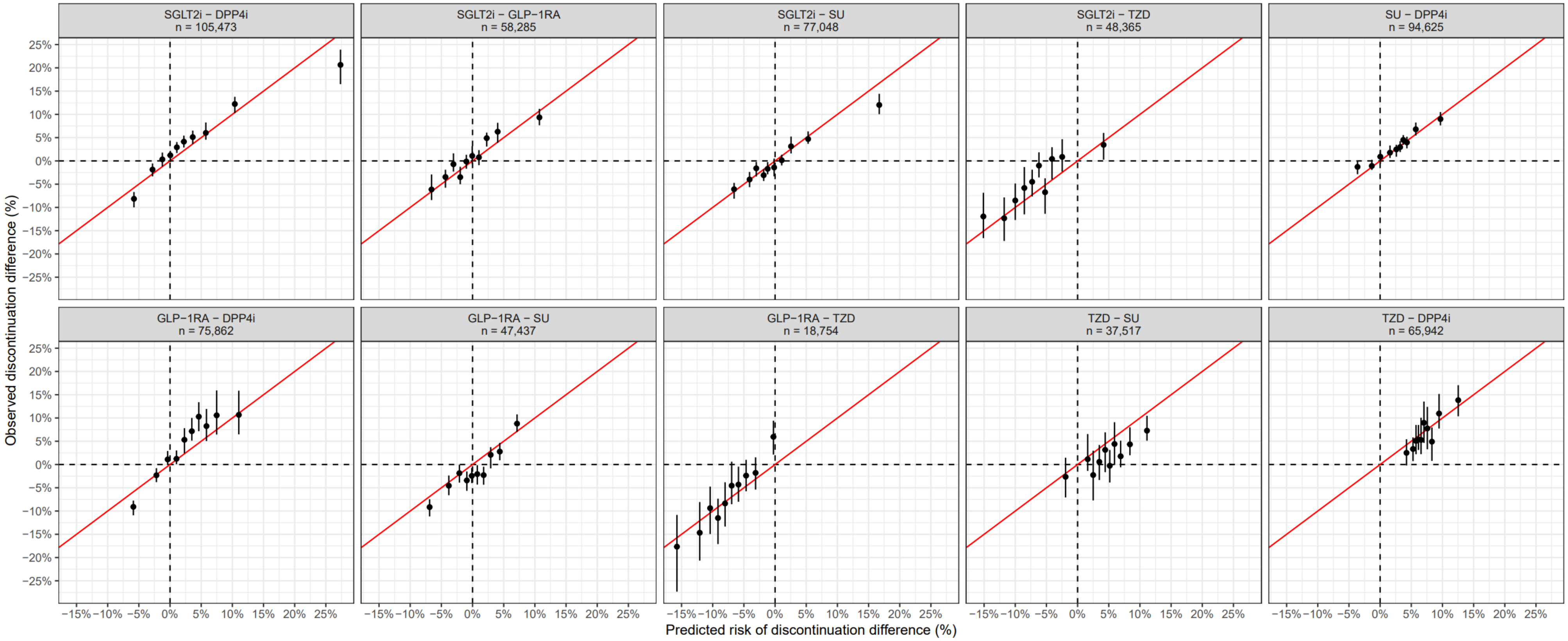

**sFig. 8: Calibration of predicted heterogeneous treatment effects across drug class pairs for 12-month discontinuation.** Red line represents perfect calibration. Point estimates represent predicted and observed differences in 12-month discontinuation. Error bars represent 95% Cis calculated through bootstrapping.

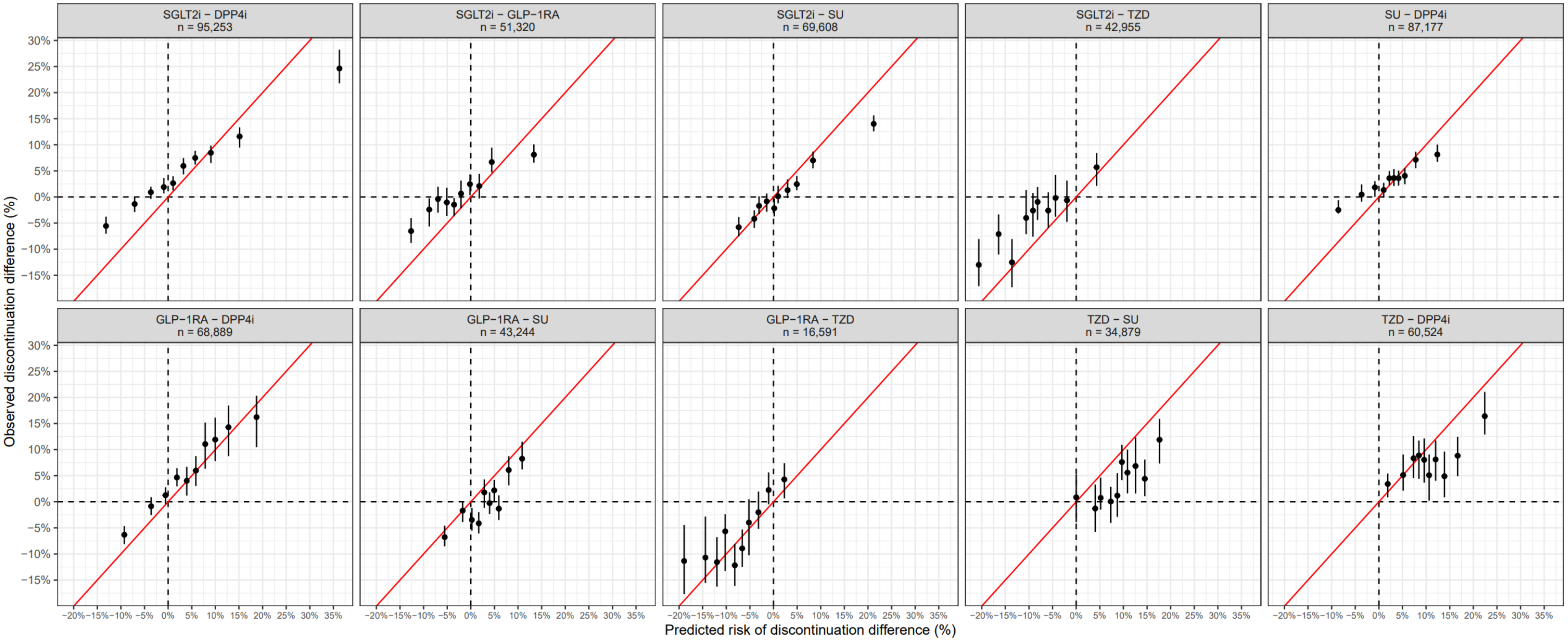

#### MASTERMIND consortium

Prof Andrew Hattersley[1], Prof Ewan Pearson[2], Dr Angus Jones[1], Dr Beverley Shields[1], Dr John Dennis[1], Dr Lauren Rodgers[1], Prof William Henley[1], Prof Timothy McDonald[1], Prof Michael Weedon[1], Prof Nicky Britten[1], Catherine Angwin[1], Dr Naveed Sattar[3], Dr Robert Lindsay[3], Prof Christopher Jennison[4], Prof Mark Walker[5], Prof Kennedy Cruickshank[6], Dr Salim Janmohamed[7], Prof Christopher Hyde[1], Prof Rury Holman[8], Prof Andrew Farmer[8], Prof Alastair Gray[8], Prof Stephen Gough[8], Dr Olorunsola Agbaje[8], Dr Trevelyan McKinley[1], Dr Sebastian Vollmer[9], Dr Bilal Mateen[7], Prof William Hamilton[1], Dr Katie G. Young[1], Dr Pedro Cardoso[1], Dr Laura Güdemann[1]

[1] University of Exeter
[2] University of Dundee
[3] University of Glasgow
[4] University of Bath
[5] University of Newcastle
[6] Kings College London
[7] University College London
[8] University of Oxford
[9] University of Kaiserslautern
